# Modelling daily infections with Covid-19 in Germany, France, and Sweden with a trend line based on day-to-day reproduction rates

**DOI:** 10.1101/2020.06.03.20121459

**Authors:** Dieter Mergel

## Abstract

The number of persons daily infected with Covid-19 as a function of time is fitted with a trend line based on an iterative power law (*n* = ¼) with a day-to-day reproduction rate modelled with a polyline. From the trend line, an effective reproduction rate *R*_eff_ of Covid-19 is calculated. In all three states, *R*_eff_ decreases in the initial phase to one indicating that there is no exponential growth. In Sweden, a steady state with *R*_eff_ around 1 and a high daily infection rates. In Germany, *R*_eff_ = 1 is reached before public and private life is restricted. With these restrictions, *R*_eff_ is reduced further to 0.87 (CI95 [0.83.; 0.91]) after 40 days so that, speculatively estimated, 9500 premature fatalities within two months may have been avoided. In France, it seems that only strongly restricting private life sends *R*_eff_ down to 1 and further down to about 0.7 (CI95 [0.3; 1.1]) after 45 days. With *R*_eff_ permanently below 1, an exponential decline of the number of daily infections is observed in Germany and France.

## 1 Introduction

Effective reproduction rates of Covid-19 are often calculated locally on a time line, e.g. by dividing numbers of daily infected averaged over 4 days by the number of daily infected 4 days before, also averaged over 4 days (an der Heiden, 2020).

In areas of automatic speech recognition (Ney and others 1992) modelling of spectra (Mergel and Qiao 2002; Qiao and others 2006; Schipporeit and Mergel 2018) and x-ray diffractograms (Davies and others 2008), integrated approaches, tackling the data set as a whole, are applied avoiding local or temporal decisions. In this paper, such an integral approach is presented by laying a trend line through the data, here daily infections, and therefrom calculating, in a second step, the effective reproduction rate taking into account the incubation process. This may better represent the chronological development of the pandemic. The trend line is based on an iterative power law with a power of ¼ applied to a hypothetical day-to-day reproduction rate.

## 2 Results and discussion

### 2.1 Investigation for Germany

The investigation for Germany is based on the number of infections as a function of the time of symptom onset, as reported to RKI (Robert Koch Institut) or imputed by RKI (Täglicher Lagebericht 2020), not the date when reported to RKI.

The data are shown in Figure 1 presenting the number *n*_Rpt_ of newly infected (dotted line, right *y*-axis) as a function of symptom onset at time *t* (in days) from March 1 (*t* = 1) to May 21, 2020 (*t* = 81), together with a trend line *n*_Calc_ (bold black line) to be explained in section 2.2. The coefficient of determination is *R*² = 0.98.

**Figure 1.**
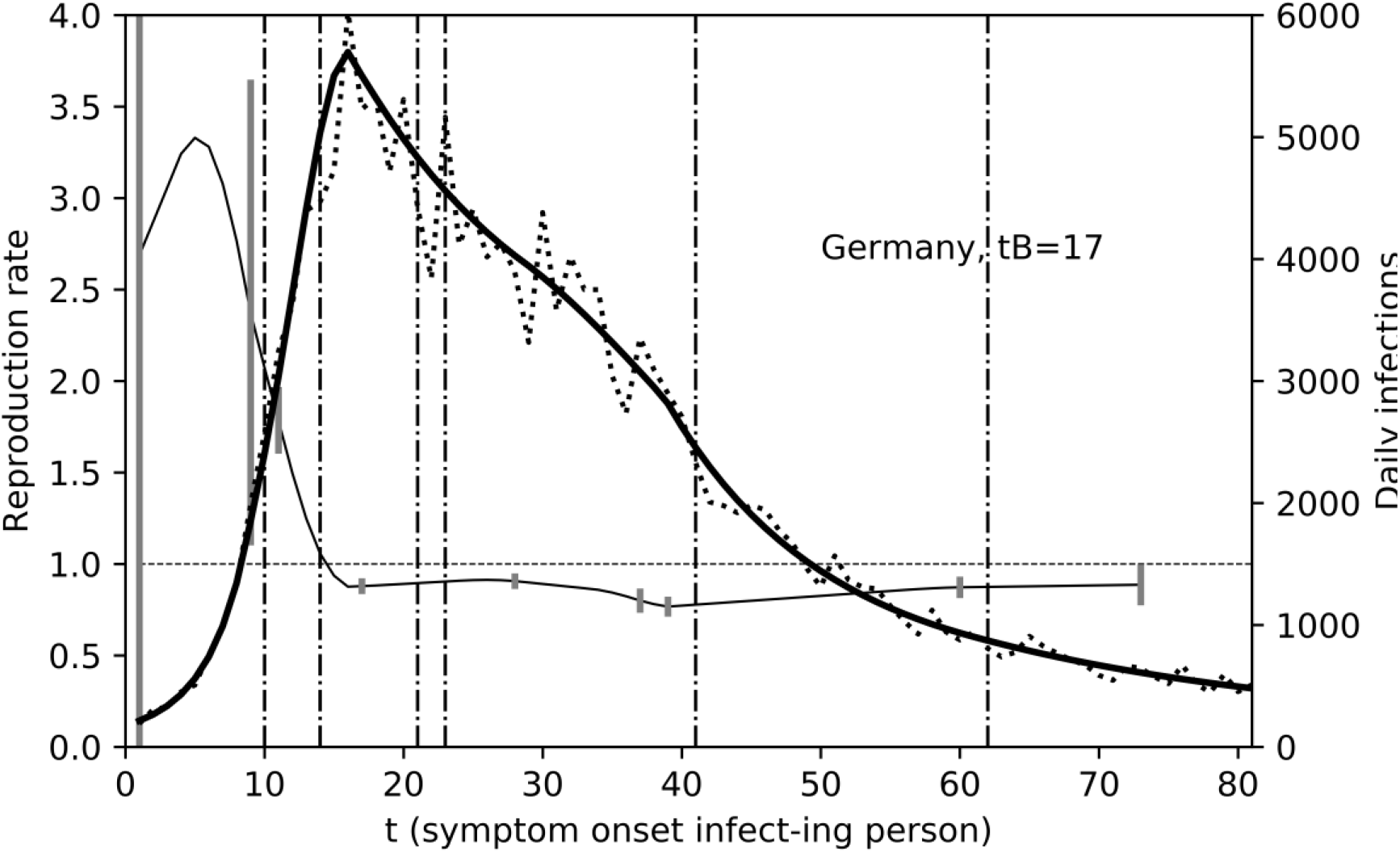
Number *n*_Rpt_ of daily infections (dotted line, right *y*-axis) reported to RKI (unsmoothed data) but presented as a function of symptom onset (an der Heiden and others 2020) together with a trend line *n*_Calc_ (bold solid line) from which the effective reproduction rate *R*_eff_ (left y-axis) is calculated.

The effective daily reproduction rate *R*_eff_ (thin black line, left *y*-axis) is calculated from the trend line as an average over three days with equal weights:

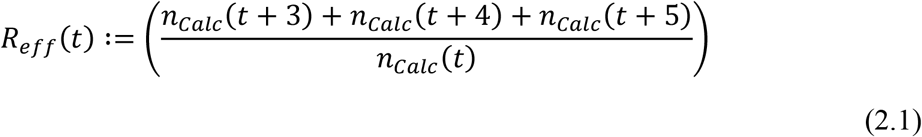

Definition (2.1) is based on (Böhmer and others 2020) where the average time between symptom onset of a person A and symptom onset of a person B infected by A is reported to be 4 on average and the median serial interval (25% to 75%) to be 3 to 5 days. The result is presented at the time of symptom onset of the infect*ing* person.

The vertical dotted lines mark political measures to reduce contacts. or social events potentially increasing contacts:

- 10.03. mass events prohibited,
- 14.03. schools closed.
- 21.03. public life restricted.
- 23.03. private contacts officially restricted,
- 10.04. Good Friday (*t* = 41), Easter weekend,
- 01.05. Weekend May 1 (*t* = 62).

The development of the reproduction rate *R*_eff_ with time is qualitatively similar to the curve published in figure 4 of (an der Heiden 2020). In Figure1, however, *R*_eff_ is plotted over the time of symptom onset of the infect*ing* person and thus better represents the definition of the effective reproduction rate and the infection process. Furthermore, calculating the effective reproduction rate with Equation (2.1) from a trend line yields better noise reductions and tends to exhibit more reliable details.

**Figure 2.**
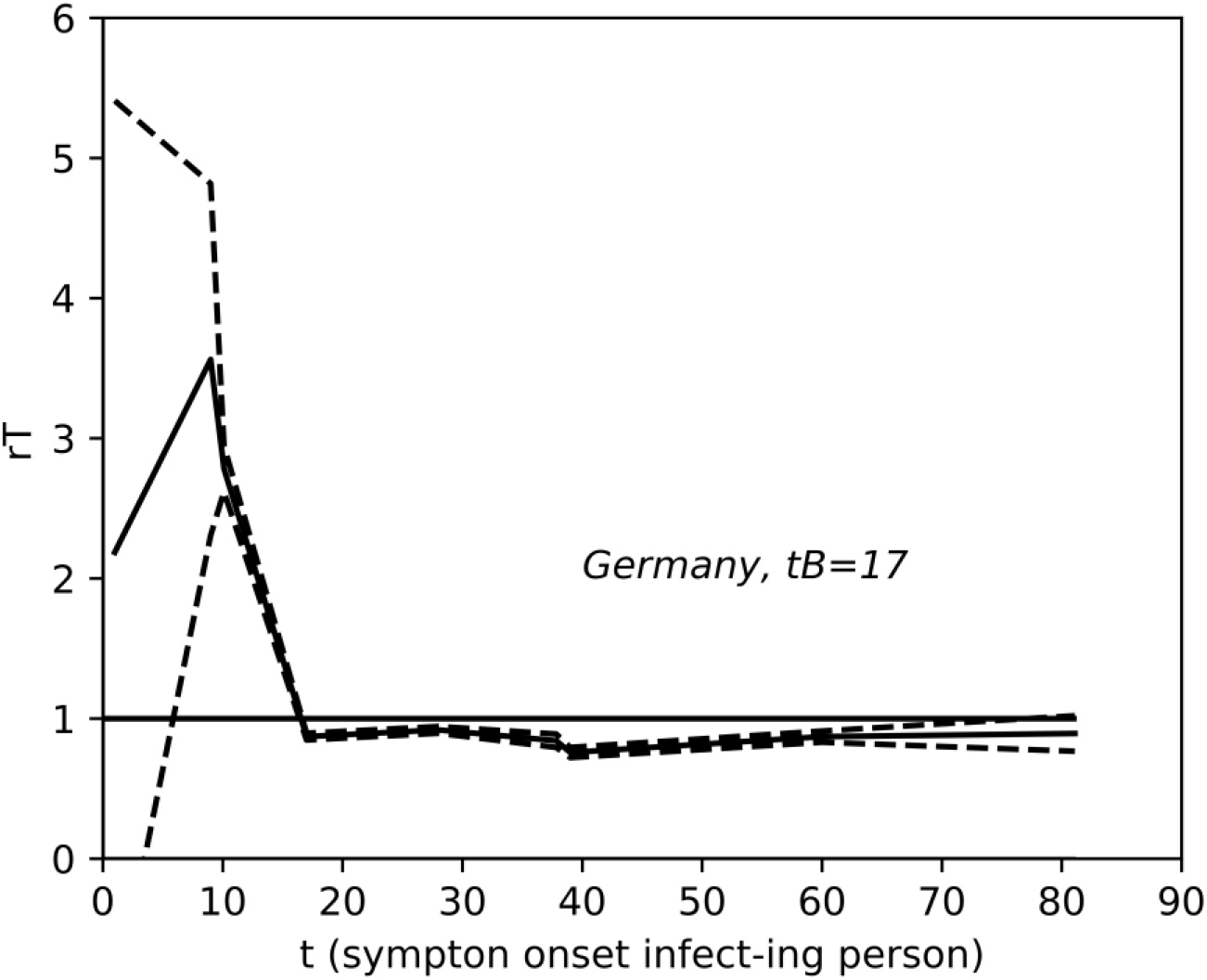
Optimized day-to-day reproduction rate *r*_T_ for Germany together with the CL95 interval as dashed lines.

The peak of *R*_eff_ in Figure 1 around *t* = 5 may be an artefact due to increasing testing in that time period, overestimating *R*_eff_.

Prohibiting mass events as of March 9 (*t* = 10), perhaps together with self-regulatory processes like voluntary reduction of contacts, sends *R*_eff_ down towards 1, before schools are closed and public life is restricted. *R*_eff_ stays below 1 after March 16. Restricted public life and private contacts decrease it in the long trend down to *R*_eff_ = 0.76 (CI95 [0.72; 0.80] as obtained from figure 2) at April 8 (*t* = 39). This leads to an exponential decrease of the number of daily reported infections down to 760 at May 3 (*t* = 64).

Exponential behavior is observed when the reproduction rate is constant over a period of time. So, exponential increase of the number of infections is not evident from the data between day 5 and 17 but well exponential decrease after day 18. The description as “exponential decrease” is justified because *R*_eff_ is approximately constant (≈ 0.8) as of day 15.

The slight temporary increase of *R*_eff_ after *t* = 20, i.e. even after restricting public and private life, may be due to spread within communities in common isolation. The effect of restricting private contacts is only seen in the long-run. This observation highlights an assumption underlying most estimates of the effective reproduction rate: the distribution of infected persons over demographic groups is supposed to remain the same in the course of time.

After the Easter weekend (*t* = 41), *R*_eff_ increases slightly again.

*R*_eff_ =1 is reached at March 14. Setting the day-to-day reproduction rate constant at 1 as of that date keeps *n*Calc constant at its maximum level 5700, hypothetically leading to 210,000 more (reported) infections for the following 67 days and about 9500 more deaths (for fatality rate 4.5% of reported infections).

### 2.2 Calculating the trend line

The equation for the trend line is based on the definition of the effective reproduction rate. When a person A with disease outbreak at time *t* infects another person B, disease outbreak for person B will be at *t* + *d*, *d* days later. Therefore, the effective reproduction rate is of the form:

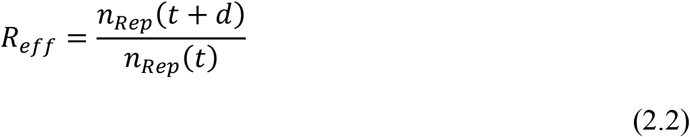

Such a formula is applied by RKI individually for every time with *d* = 4 and suitable averaging over 4 days (an der Heiden 2020).

For our trend line, we choose an iterative power law with a hypothetical “day-to-day reproduction rate” *r_T_*(*t*)

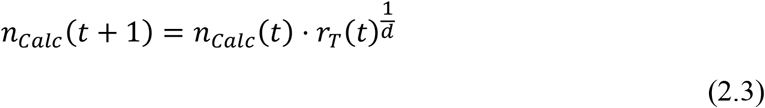

For constant *r_T_*, we get *n_Calc_*(*t* + *d*) = *n_Calc_*(*t*) · *r_T_*, then *R*_eff_ = *r*_T_ and an exponential increase/decrease of the number of daily infected is obtained. So, our approach may be characterized as exponential growth with a base changing over time.

Principally, *d* is a fit parameter, but we fix *d* = 4 to come close to the reported incubation time and Equation (2.1). Power ¼ is equivalent to the 4th root.

The day-to-day infection rate can also be calculated backwards:

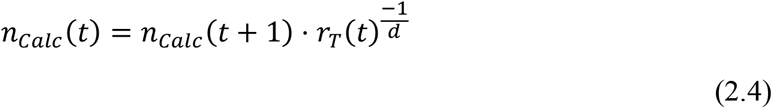

so that we may conveniently choose a border time *t*_B_ to calculate with Equation (2.3) above *t*_B_ and with Equation (2.4) below *t*_B_ avoiding starting with a very low number at *t* = 1.

The daily reproduction rate *r*_T_(*t*) as a function of time is modelled as a polyline with vertices (*t*_V_*, r*_V_) with the first and last *t*_V_ being begin and end of the data series (here 1 and 81). The inner values of *t*_V_ and all values of *r*_V_ are optimized with a solver algorithm (*Excel solver* (Mergel 2017; Schipporeit and Mergel 2018) to yield the best least-square fit of the trend line *n*_Calc_ to the reported data *n*_Rep_. The data for Germany are modelled with 9 vertices yielding the optimized day-to-day reproduction rate *r*_T_ in Figure 2.

As solver algorithms do not guarantee a global minimum, some heuristics is needed to make an initial guess of the fit parameters. The border time *t*_B_ is chosen close to the maximum. The reproduction rate around the maximum is about 1, for increasing number of infections, i.e. before the maximum, *R*_eff_ is greater than 1, for decreasing number of infections it is smaller than 1. When the curve changes rapidly, more vertices are needed. With these guidelines, the vertices are chosen manually within an Excel spreadsheet until the trend line lies approximately within the reported data and solver can do the rest.

In order to get confidence intervals of the *r*_V_, the calculation is reproduced within *Python* with the function *curve_fit* from the *scipy.optimize* library. This function outputs the covariance matrix from which the standard error of the fit parameters can be calculated as square root of the diagonal elements. The degree of freedom for the fit is 68, obtained as 84 data points minus 16 fit parameters, giving a Student’s *t*-value of 2.0 for a confidence level of 95%. The CL95 confidence interval is, therefore, calculated as ± 2.0·standard error and shown in Figure 2 with dashed lines. The CI95 intervals are [2.6; 2.9] at around *t* =10 and [0.72; 0.8] at around *t* = 39 (April 8).

The polyline *r*_T_(*t*) is only used to obtain the best trend line for the daily infections and does not enter the determination of the effective reproduction rate directly.

The more fluctuating data for France and Sweden are modelled with 8 vertices. Student’s *t* is 2.0 for France (degree of freedom is 73 - 14) and 1.99 for Sweden (degree of freedom 90 - 14).

### 2.3 Investigation for Sweden

The trend line for the day-to-day reproduction rate is based on only 7 vertices (*t*_V_, *r*_V_) with the positions *t*_V_ a-priori chosen (“informed guess”) and only the *r*_V_ varied.

Figure 3 shows the number of daily reported infections in Sweden as a function of the report time together with the trend line and the effective reproduction rate calculated from the trend line. The vertical bars indicate the 95% confidence intervals calculated from the corresponding values for *r*_T_. At *t*= 63, *R*_eff_ =0.98, CI95 = [0.93; 1.03].

**Figure 3.**
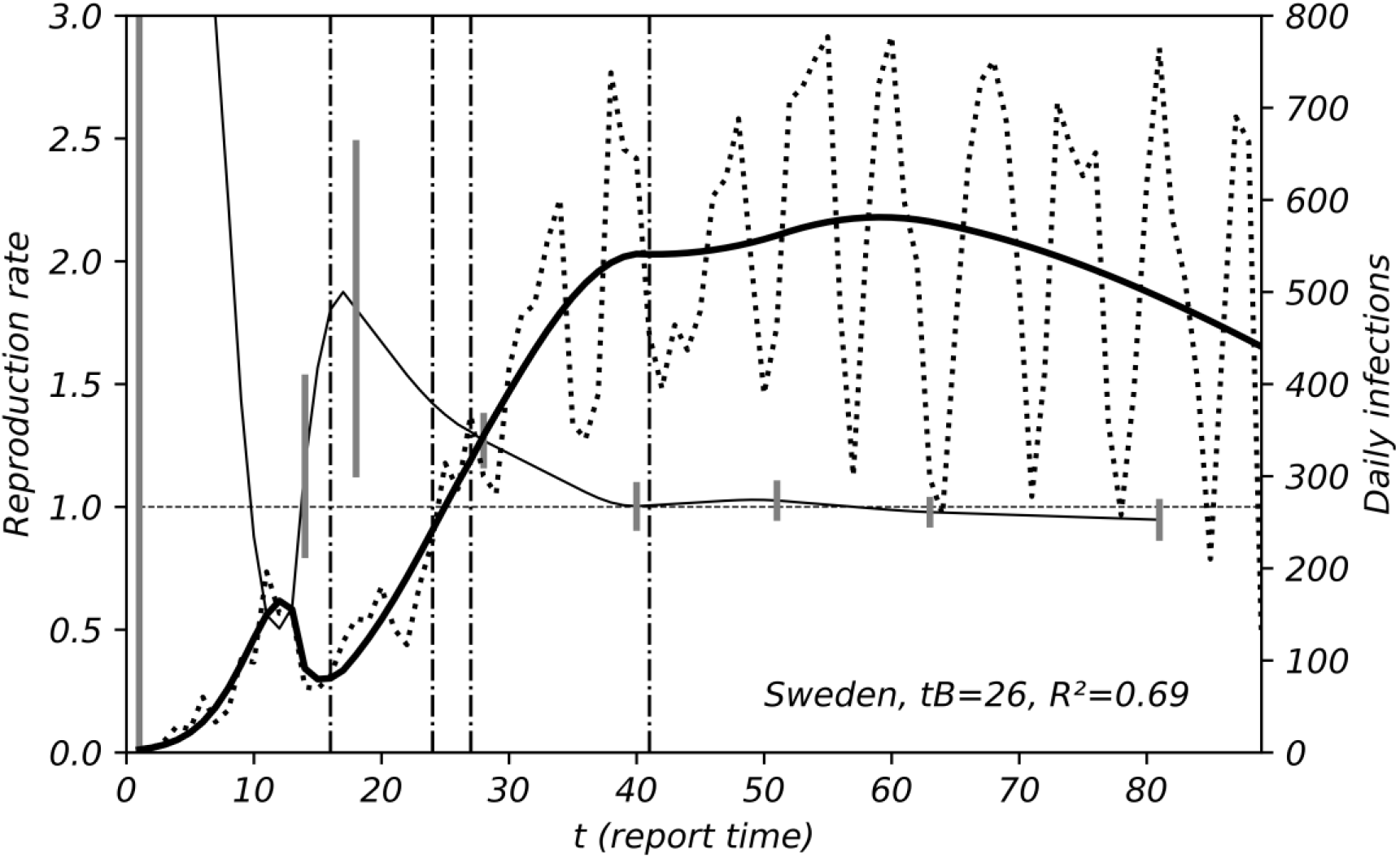
Daily infections *n*_Rpt_ in Sweden (dotted line, right *y*-axis) reported to the Swedish Health Agency (Folkhälsömyndigheten), as a function of reporting time, up to May 28 (*t* = 91) together with a trend line (bold solid line); effective reproduction rate calculated from the trend line (thin solid line, left *y*-axis)

The vertical dashed lines indicate dates of rules to restrict social contact or social events potentially increasing contacts:

- 16.03. people elder than 70 are advised to stay home,
- 24.03. bar openings restricted,
- 27.03. gatherings with more than 49 people prohibited,
- 10.04. Good Friday, Easter weekend.

The “soft” measures of the Swedish administration sends *R*_eff_ down to about 1 in the long run, leading to a constant number of newly infected between day 40 and day 60 in Figure 3. The decrease after *d* = 60 might be due to beginning weakening of the infection process.

The residuals *n*_Rpt_-*n*_Calc_ of the fit are presented in Figure 4 together with the function: (2.5)

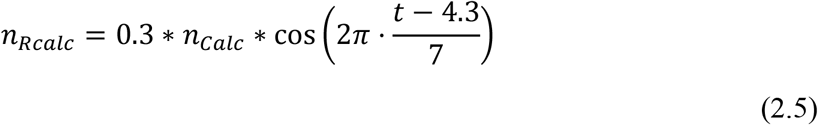

intended to model weekly fluctuations. The coefficient of determination is *R*²= 0.58.

**Figure 4.**
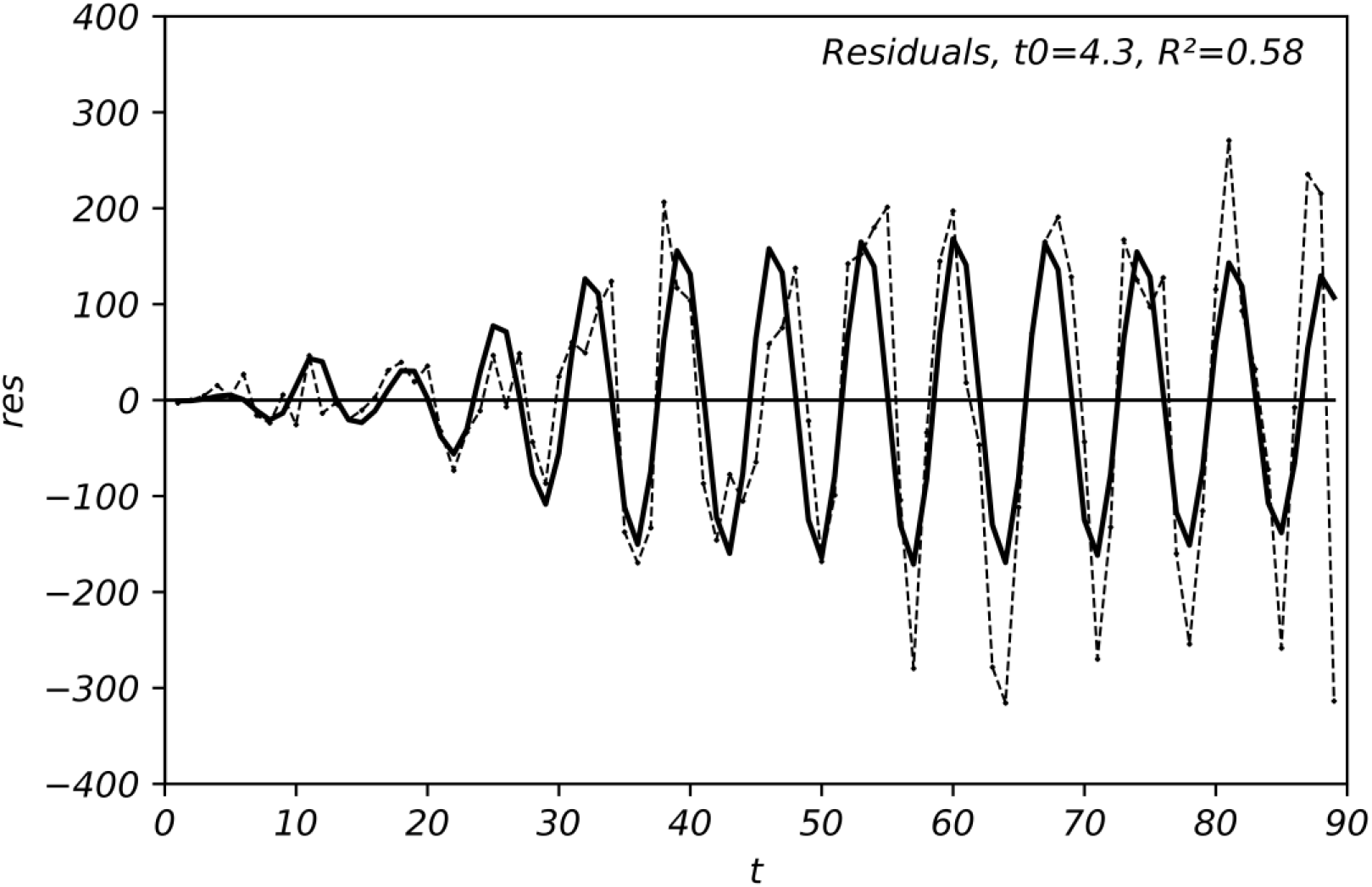
Residuals *n*_Rpt_ – *n*_Calc_ of the data from Sweden as a function of report time together with a cosine fit function

As the residuals *n*_Rpt_-*n*_Calc_ are systematically described by equation (2.5), we recalculate *R*² of the fit in Figure 3 with the variance of *n*_Rpt_-*n*_Calc_ as residual variance and the variance of the reported daily infections as total variance, the result being *R*² = 0.87. This *R*² is claimed for the trend line in Figure 3.

Equation (2.5) has its maximum between the 4^th^ and the 5^th^ day after Saturdays (t = 0 is Saturday, February 29) so that it is tempting to attribute the increased number of cases not only to the reporting process but also to weekends meetings.

### 2.4 Investigation for France

The number of newly infected *n*_Rep_ in France as reported to the Agence régionale de santé (ARS 2020) is shown in Figure 5 together with the trend line and the effective reproduction rate calculated therefrom. The data point at May 5 (*t* = 64), presented with a diamond, is regarded an outlier and replaced for the calculation by the average over neighboring values. The coefficient of determination is *R*² = 0.76. Trying to describe the fluctuations as a weekly periodic process as for Sweden, potentially increasing *R*², is not successful.

**Figure 5.**
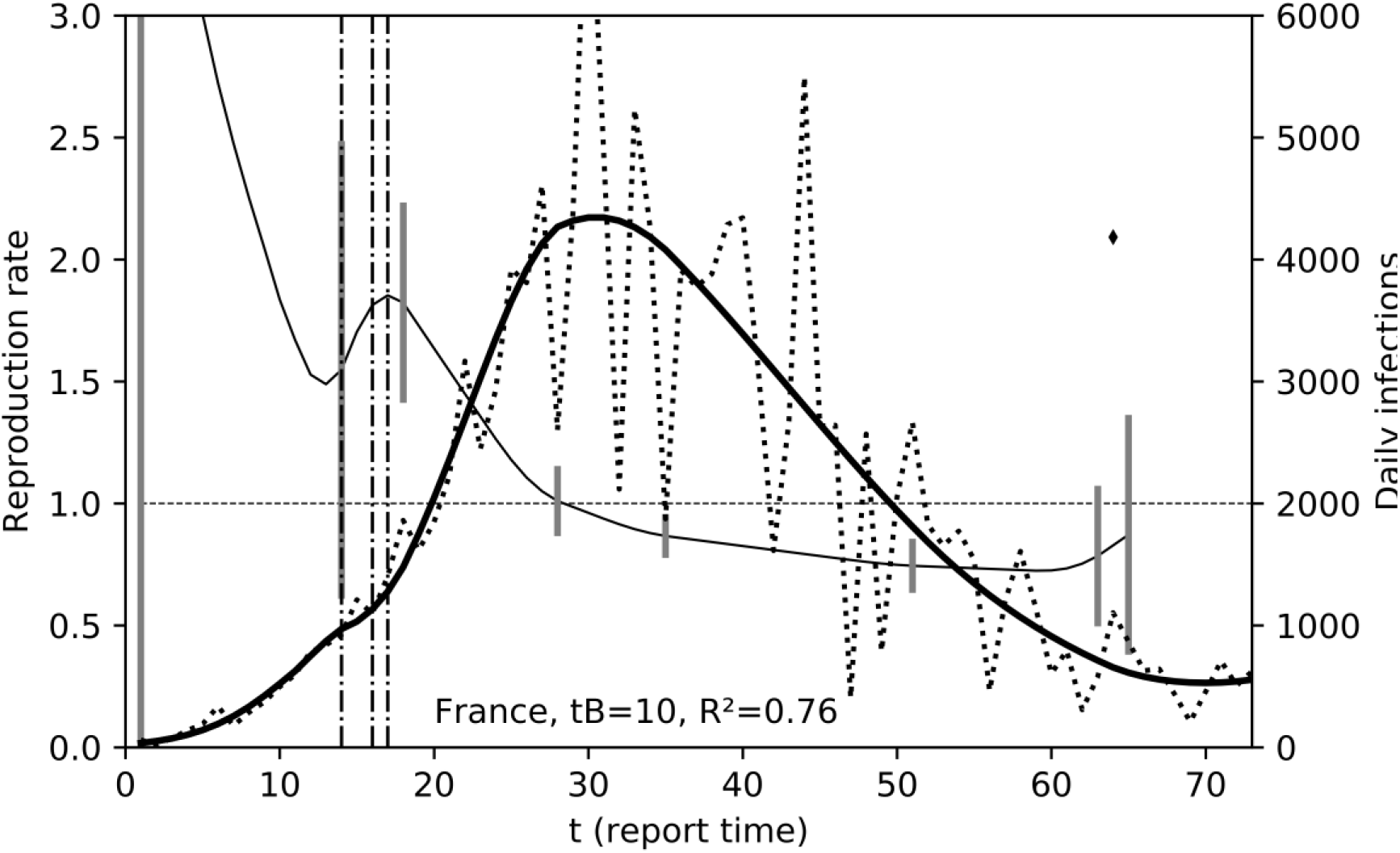
Number of newly infected in France (dotted line, right *y*-axis) as a function of the time reported to ARS (ARS 2020) together with a trend line *n*_Calc_ (bold solid line); the data point marked with a diamond is regarded an outlier; effective reproduction rate calculated from the trend line (thin solid line, left *y*-axis)

The vertical dotted lines represent the dates of restricting measures:

- 14.03. closing public institutions
- 16.03. schools closed
- 17.03. confinement ordered (only 1 h leave every day)

These measures send down *R*_eff_ to about 0.75 (CI95 [0.65; 0.85]) at *t* = 51.

## 3 Summary and conclusions

A trend line based on a day-to-day reproduction rate is able to deliver visually reasonable fits to the number of daily infections. The effective reproduction rate is calculated from this trend line in a second step assuming incubation times between 3 to 5 days and is plotted over the relevant time of the infect*ing* person. The degree of freedom is more than 60 for the three countries. This model is claimed to make the best use of the available information.

For an interpretation of the results it must be assumed that the reported infections always represent the infection process along the timeline in the same way. This is not the case when the number of tests is changed or the test limit is reached. Another problem is when the demographic distribution of the infections changes over time. However, taking the numbers at face value we draw the following hypothetical conclusions that we would like to bring up to discussion.

1. Exponential *increase* of the number of daily infections is not observed, as the effective reproduction rate decreases after a statistically uncertain initial phase and reaches a value of 1 after some time for all three countries.
2. Exponential *decrease* is observed for Germany and France after lockdown measures keeping *R*_eff_ constantly below 1 (≈ 0.8).
3. For Sweden without stricter lockdown measures, *R*_eff_ stays at around 1 until after 60 days the infection process seems to weaken. The number of new daily infections remains stationary but high at about 500 (of 10 million). The weekly periodic variation of new daily infections might indicate relatively more infections during weekends.
4. In France, severe shutdown measures seem to have been necessary to bring *R*_eff_ to below 1 and daily infections down to about 600 (of 67 million).
5. It remains to be seen whether similar measures as in Sweden after relaxing shutdown could keep the current daily infection rate stationary also in Germany and France.
6. By setting the day-to-day reproduction rate constant to 1 after it is reached for the first time as is the case for Sweden, the number of infections stays at its maximum and in Germany 210,000 more infections are to be expected within 65 days. So, by restricting public life and private contacts, about 9500 premature deaths may have been avoided within that period of about two months.

## Data Availability

https://www.statista.com/statistics/1102193/coronavirus cases development in sweden/
an der Heiden, M., Hamouda, O. (2020): Schaetzung der aktuellen Entwicklung der SARS CoV 2 Epidemie in Deutschland Nowcasting, Epid Bull 2020;17:10-16|DOI 10.25646/6692.4
ARS / Santé Publique France / Ministere des Solidarites/Sante, Nombre de nouveau cas quotidiens de Covid-19 confirmés, 14 mai 2020

